# Statistical challenges of investigating a disease with a complex diagnosis

**DOI:** 10.1101/2021.03.19.21253905

**Authors:** João Malato, Luís Graça, Luís Nacul, Eliana Lacerda, Nuno Sepúlveda

**Affiliations:** Instituto de Medicina Molecular João Lobo Antunes, Faculdade de Medicina, Universidade de Lisboa, Portugal; Department of Clinical Research, Faculty of Infectious and Tropical Diseases, London School of Hygiene and Tropical Medicine, United Kingdom; Department of Infection Biology, Faculty of Infectious and Tropical Diseases, London School of Hygiene and Tropical Medicine, United Kingdom; Centro de Estatística e Aplicações, Universidade de Lisboa, Portugal

**Keywords:** multimensional scaling, cluster analysis, misclassification, cohen’s *κ* coefficient, Jaccard’s similarity index

## Abstract

Given the absence of a disease-specific biomarker, there are more than 20 symptoms-based case definitions of myalgic encephalomyelitis/chronic fatigue syndrome. As a consequence, the diagnosis for a given patient could vary from one case definition to another. In this context, we analyse data from a biobank dedicated to this disease in order to study the agreement between different case definitions, the similarity between symptom’s profile among all participants including healthy controls and patients with multiple sclerosis. We also investigate the impact of patients’ misclassification on a hypothetical association analysis using data simulation.

## 1 Introduction

Myalgic encephalomyelitis/chronic fatigue syndrome (ME/CFS) is a complex disease whose patients manifest unexplained fatigue lasting for more than six months (Fukuda et al., 1994) or suffer from post-exertional malaise that is not alleviated by rest (Carruthers et al., 2003). Disease prevalence has been estimated between 0.4% and 1.0% affecting six women to one man (Morris and Maes, 2013). The underlying pathological mechanisms remain poorly understood, but they are often associated with environmental stressors, including severe viral infections (Rasa et al., 2018).

Until now there is no accurate biomarker for disease diagnosis. To overcome this problem, researchers and clinicians altogether have proposed more than 20 different case definitions based on patients’ symptomatology while excluding known diseases that could explain the fatigue reported by suspected cases (Brurberg et al., 2014). As a consequence, the diagnosis for a given patient can vary from one case definition to another. Therefore, research from ME/CFS could be affected by the inclusion of false positive cases in the respective data.

In the present paper, we discuss the problem of diagnosing ME/CFS using data from the United Kingdom ME/CFS Biobank (UKMEB). With this purpose, we first introduce the biobank and its data. We then assess the agreement between 4 common case definitions of ME/CFS in 275 suspected cases belonging to the UKMEB. We then estimate the similarity between symptom’s severity profiles from suspected cases, patients with multiple sclerosis, and healthy controls. We also study the impact of patients’ misclassification on the statistical power of a hypothetical association analysis. Finally, we conclude this paper with some final remarks.

## 2 The UKMEB

The UKMEB refers to a large data set of suspected cases of ME/CFS, healthy controls, and patients with multiple sclerosis included as an additional control group (Lacerda et al., 2018). In terms of recruitment, suspected cases were identified in different institutions across the National Health Service from the United Kingdom and then referred to the CureMe group, a dedicated clinical research team based in the London School of Hygiene & Tropical Medicine (LSHTM) and responsible for recruiting, managing, and curating the biobank. For this paper, the data set under analysis consists of a total of 523 participants divided into 275 suspected cases of ME/CFS, 136 healthy controls, and 112 patients with multiple sclerosis.

## 3 Diagnostic agreement analysis

After patients’ referral for a possible integration in the biobank, suspected cases were comprehensively evaluated according to four case definitions of ME/CFS: Centre for Disease Control criteria (CDC-1994) (Fukuda et al., 1994), Canadian Consensus Criteria (CCC-2003) (Carruthers et al., 2003), Institute of Medicine Criteria (IOM-2005) (Institute of Medicine (US), 2015), and International Consensus Criteria (ICC-2011) (Carruthers et al., 2011). The CDC-1994 requires the patients to have unexplained fatigue for at least 6 months and at least four out of eight fatigue-related symptoms. The IOM-2005 is typically used by general practitioners and it requires the patients to show at least three main symptoms such as profound fatigue, post-exertional malaise, and unrefreshing sleep. The CCC-2003 requires the patients to manifest four or more fatigue specific symptoms, at least two neurological or cognitive ones, and at least one autoimmune, neuroendocrine, or immune symptom. Finally, the ICC-2011 is more focused on neuro-immune and cognitive symptoms, and on the inability to produce sufficient energy on demand (post-exertional neuroimmune exhaustion).

There were 269 (97.8%), 233 (84.7%), 229 (83.3%) and 213 (77.5%) out of 275 suspected cases whose symptoms agreed with CDC-1994, IOM-2005, CCC-2003, and ICC-2011, respectively (Table 1). This finding suggests that the general practitioners who referred the suspected cases to a possible integration in the biobank made their diagnosis based on the CDC-1994. Unsurprisingly, only 62.9% of the suspected cases (*n* = 173) had a positive diagnosis across all the four case definitions. Therefore, the remaining suspected cases had at least one negative diagnosis.

**Table 1:** Frequency of suspected cases of ME/CFS according to their diagnostic outcomes using different case definitions. Percentages in the last row indicate the proportion of diagnosed cases by each case definition.

| Case definition |  |  |  | N | % of total<br>suspected cases |
| --- | --- | --- | --- | --- | --- |
| CDC-1994 | IOM-2005 | CCC-2003 | ICC-2011 |  |  |
| + | + | + | + | 173 | 62.9 |
| + | + | + | – | 32 | 11.6 |
| + | – | + | + | 16 | 5.8 |
| + | + | – | + | 16 | 5.8 |
| + | – | – | – | 14 | 5.1 |
| + | + | – | – | 10 | 3.6 |
| + | – | + | – | 5 | 1.8 |
| + | – | – | + | 3 | 1.1 |
| – | – | + | + | 3 | 1.1 |
| – | – | – | + | 1 | 0.4 |
| – | + | – | – | 1 | 0.4 |
| – | + | – | + | 1 | 0.4 |
| 97.8% | 84.7% | 83.3% | 77.5% | 275 | 100% |

It is worth noting that there were no suspected cases who had a negative diagnosis across all case definitions. There were also three individuals whose symptoms agreed with ICC-2011 only, IOM-2005 only, or both criteria. These individuals were considered to be fatigued but non-ME/CFS patients given that they did not agree with either the CDC-1994 or the CCC-2003 as recommended for ME/CFS research (Pheby et al., 2020).

To better understand the agreement between diagnostic outcomes obtained from different case definitions, we used the Jaccard’s similarity index, *J* (Gower and Warrens, 2014). Note that this index is usually a measure used to compare objects with shared attributes. Here we instead applied this index to compare attributes themselves. For a pair of case definitions (*C*_*i*_, *C*_*j*_), this index was estimated as

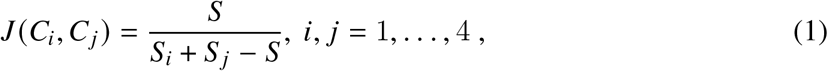

where *S*_*i*_ and *S*_*j*_ are the number of suspected cases with a positive diagnosis by *C*_*i*_ and *C*_*j*_, respectively, and *S* is the number of suspected cases with a positive diagnosis by both criteria. In theory, the index is defined between 0 and 1 (i.e., no and full agreement between *C*_*i*_ and *C*_*j*_ across all individuals, respectively).

The estimates of this index ranged from 0.752 (IOM-2005 versus ICC-2011) to 0.876 (CDC-1994 versus IOM-2005; CDC-1994 versus CCC-2003) (Table 2). The estimates showed the stringency and differences in scope of each case definition. In addition, these estimates showed that, even if the general practitioners applied two different case definitions of ME/CFS in their diagnosis, there could still be a fraction of suspected cases where the respective diagnostic outcomes might not agree with each other.

**Table 2:** Estimates of the Jaccard’s similarity index for the four case definitions of ME/CFS using data from the UKMEB.

|  | CDC-1994 | IOM-2005 | CCC-2003 | ICC-2011 |
| --- | --- | --- | --- | --- |
| CDC-1994 | 1.000 | 0.876 | 0.876 | 0.760 |
| IOM-2005 | 0.876 | 1.000 | 0.840 | 0.752 |
| CCC-2003 | 0.876 | 0.840 | 1.000 | 0.753 |
| ICC-2011 | 0.760 | 0.752 | 0.753 | 1.000 |

## 4 Symptoms’ similarity analysis

A major advantage of using data from the UKMEB is the comprehensive symptom’s characterisation of all study participants. In particular, each participant had to report the severity of 57 symptoms occurred a month before data collection. Severity of each symptom was categorised into absence, mild, moderate, and severe. These invaluable data were then analysed to assess the similarity of all participants in terms of their symptom’s severity profile. With this purpose, we first computed all possible 4 × 4 contingency tables resulting from cross-tabulating the symptom’s severity data for any given pair of participants (*i, j*), *i, j* = 1, …, 523. We then calculated a similarity matrix between any given pair of individuals by estimating the Cohen’s *κ* coefficient (Agresti, 2002) in the corresponding 4 × 4 contingency tables, that is,

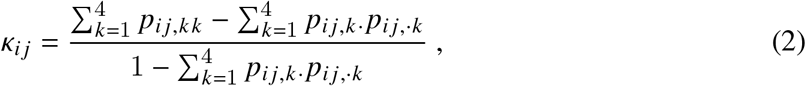

where *k* = 1, …, 4, *p*_*ij, kk*_ is the proportion of symptoms with severity *k* reported by both individuals *i* and, *p*_*i j,k*·_ is the proportion of symptoms with severity *k* reported by individual *i*, and *p*_*i j*, ·*k*_ is the proportion of symptoms with severity *k* reported by individual *j*. The resulting similarity matrix was then analysed by classical multidimensional scaling (MDS) (Figure 1A) and hierarchical cluster analysis using complete linkage (Figure 1B).

**Figure 1:**
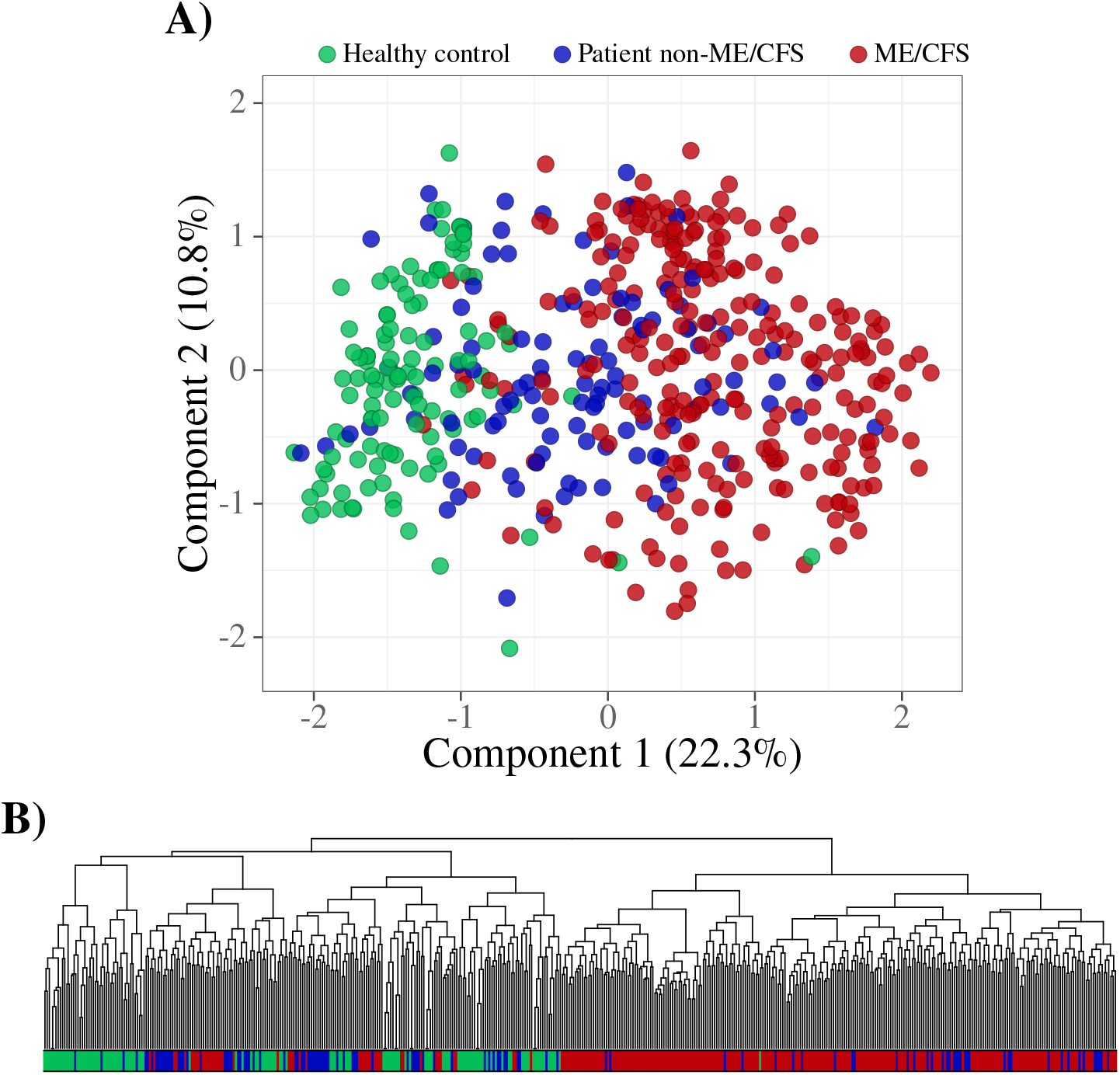
Symptom’s similarity analysis based on the Cohen’s *κ* coefficient: classical multidimensional scaling (A); dendrogram of hierarchical clustering analysis based on complete linkage (B) where the colour coding at the bottom is the same shown in A.

With respect to the classical MDS, the first two components could explain 33.1% of the total inertia (Figure 1A). More importantly, the first component clearly discriminated healthy controls from suspected cases of ME/CFS. In the same component, patients with multiple sclerosis and the three fatigued non-ME/CFS cases were located between these two groups with some overlap. As expected, healthy participants were the most homogeneous cohort due to an absence or, at most, mild severity of the different symptoms. In contrast, the suspected cases of ME/CFS consisted of a diverse group as evidenced by their wide spread in the plot. Interestingly, a few suspected cases of ME/CFS had symptom’s severity profiles similar to the ones from healthy controls. In agreement with these observations, the hierarchical cluster analysis revealed that some suspected cases of ME/CFS could be placed in clusters together with healthy controls and patients with multiple sclerosis (Figure 1B); a detailed analysis on the optimal number of clusters will be done elsewhere. Therefore, it was reasonable to assume that some of the suspected cases of ME/CFS, although agreeing with CDC-1994 or CCC-2003, could be in fact true cases of another disease, as discussed by Nacul et al. (2017).

## 5 Impact of misclassification on an association analysis

Given the possibility of patients’ misclassification, we performed a small simulation study to assess the reduction of statistical power attributed to this issue in the context of an association analysis. With this purpose, we simulated data from a case-control study with the aim to investigate a hypothetical association of a binary exposure variable (exposed versus not exposed) with ME/CFS. In this scenario, the observable data could be summarised by a 2×2 frequency table whose sampling distribution was given by the following product of two Binomial distributions,

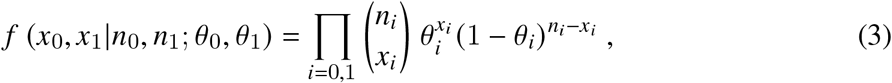

where *x*_0_ and *x*_1_ are the frequencies of exposed healthy controls and suspected cases, respectively, *n*_0_ and *n*_1_ are the associated sample sizes, and *θ*_0_ and *θ*_1_ are the corresponding probabilities of exposure in healthy controls and suspected cases.

To study the impact of a potential misclassification of suspected cases on the detection of a possible association, four main assumptions were considered for the simulated data: (i) suspected cases could be divided into apparent (or false positive) cases and true positive cases of ME/CFS; (ii) the apparent cases were deemed equivalent to healthy controls in terms of degree of exposure, i.e., the probability of exposure in these individuals was given by *θ*_0_; (iii) there was an overall misclassification rate, *γ*, for the suspected cases; and (iv) misclassification was only dependent on the true clinical status of each suspected case. Under the assumption (ii) and the law of total probability, the probability of exposure associated with suspected cases could be written as

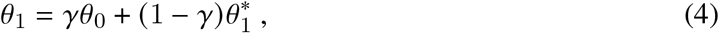

where 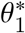 is the probability of exposed true cases.

We then studied the power of rejecting the null hypothesis of lack of association (i.e., *H*_0_: odds ratio = 1) by the Pearson’s *χ*^2^ test for independence, when considering this simple misclassification scenario. Similar investigation could have been done using Fisher’s exact test instead. With this purpose, we used simulation to estimate the number of times that *H*_0_ could be rejected at a significance level of 5%.

We augmented the observable 2×2 frequency table where the suspected cases were subdivided into apparent and true positive cases (Table 3). In this case, we simulated data from healthy controls according to the Binomial distribution with a sample size of *n*_0_ individuals and probability of success *θ*_0_. With respect to the suspected cases, we simulated data from a Multinomial distribution with a sample size of *n*_1_ individuals and probability vector given by the probabilities shown in Table 3. Note that, given assumption (iv), the associated Multinomial distribution could be decomposed into the following Binomial distribution

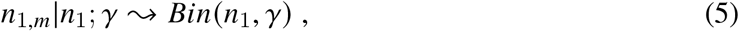

referring to how many individuals were hypothetically misclassified as true positive cases, and two Binomial distributions conditional to *n*_1,*m*_

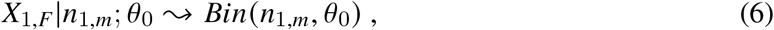

and

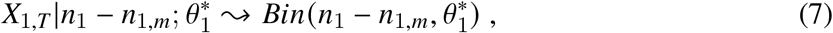

where *X*_1,*F*_ and *X*_1,*T*_ were the random variables referring to the number of exposed false positive and true positive cases, respectively.

**Table 3:**
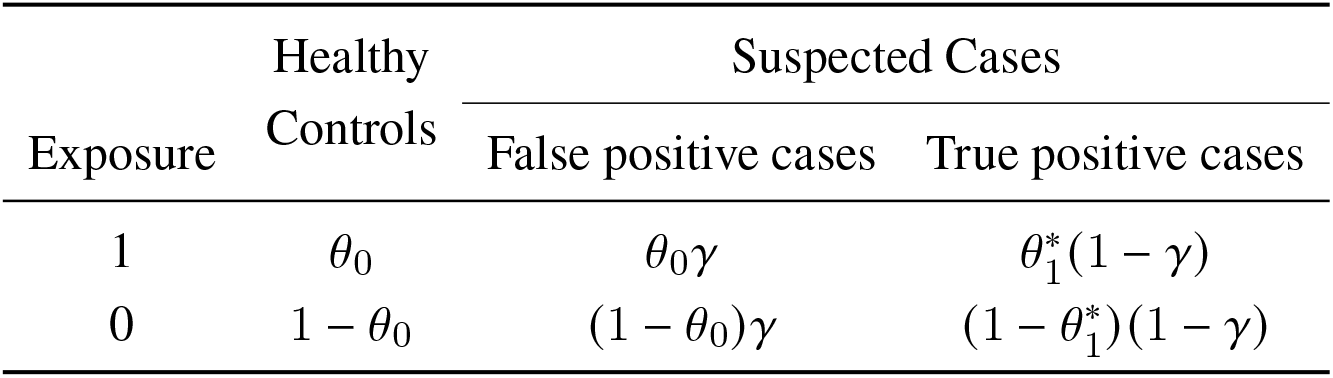
Augmented version of the observable 2 × 2 frequency table and the respective probabilities under a Binomial and a Multinomial distribution for healthy controls and suspected cases, respectively.

| Exposure | Healthy Controls | Suspected Cases |  |
| --- | --- | --- | --- |
|  |  | False positive cases | True positive cases |
| 1 | $\theta_0$ | $\theta_0\gamma$ | $\theta_1^*(1 - \gamma)$ |
| 0 | $1 - \theta_0$ | $(1 - \theta_0)\gamma$ | $(1 - \theta_1^*)(1 - \gamma)$ |

For illustrative purposes, we performed our simulation study with *n*_0_ = *n*_1_ = 100, *θ* _0_ = 0.25, and 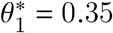. According to this parameter specification, the odds ratio of true positive cases versus healthy controls was 1.62, a low but reasonable value for a putative association with ME/CFS, given that there is no disease-specific biomarker. To estimate the power of rejecting *H*_0_, we generated 10,000 data sets for each value of *γ*, ranging from 0 (no misclassification) to 1 (full misclassification) with a lag of 0.01. In each data set, *H*_0_ was rejected if the p-value of the Pearson’s *χ*^2^ test was less than 0.05. For a given parameter set, power was finally estimated as the proportion of simulated data sets in which *H*_0_ was rejected.

As expected, the estimated power decreased with the misclassification rate *γ* (Figure 2). As a control scenario, when all suspected cases were considered to be false positives (*γ* = 1) and therefore the data sets were simulated from *H*_0_, the corresponding power was estimated at 5%, the significance level specified for the Pearson’s *x*^2^ test. In opposition, when the suspected cases were all considered true positive cases (*γ* = 0), the power to detect a hypothetical association was estimated at 34%. This low power simply reflected the limited sample size to detect a weak association between exposure and the disease. In a less extreme case of misclassification, *γ* = 10% implied an estimated power of 29%, which reflected a decrease in 14.7% of the power estimated for the scenario with no misclassification.

**Figure 2:**
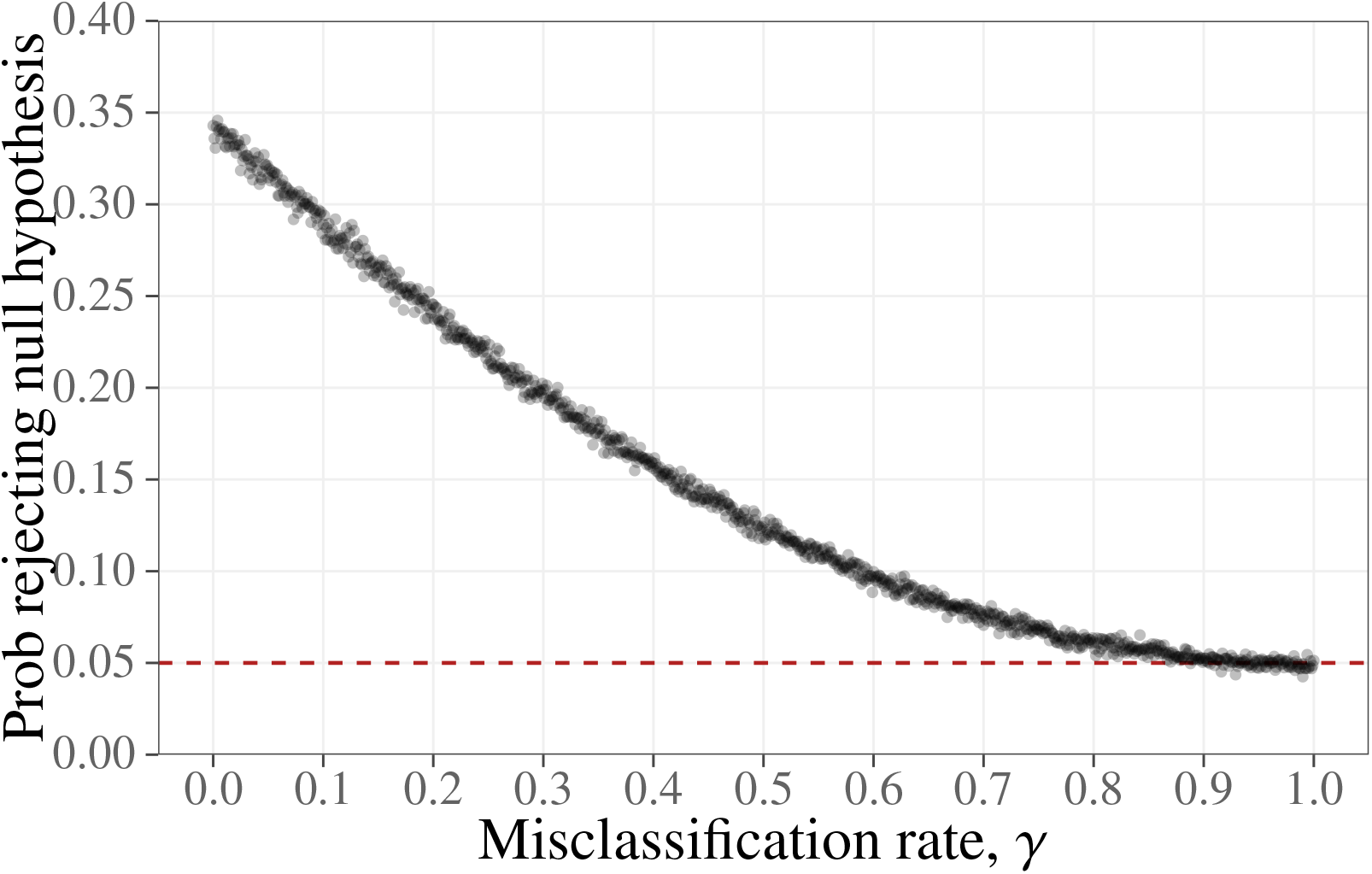
Estimated probability of rejecting *H*_0_ (i.e., lack of association) as function of the misclassification rate *γ*.

## 6 Concluding remarks

In summary, our analysis showed that suspected cases of ME/CFS from the UKMEB did not fully agree with four main case definitions of the disease. In addition, some of these suspected cases showed symptom’s severity profiles similar to healthy controls and patients with multiple sclerosis. These findings demonstrated the difficulty of diagnosing ME/CFS based on symptoms’ assessment alone. To overcome this and other difficulties, there are currently efforts for a stronger collaboration among European researchers for accelerating the discovery of an objective disease-specific biomarker (Scheibenbogen et al., 2017). However, joint efforts for biomarker discovery are very likely to suffer from limited statistical power due to a possible misclassification of the suspected cases. A possible solution to this problem is to take into account for misclassification in the respective statistical analysis. Such a solution is also problematic because modelling misclassification leads to an eventual problem of overparameterisation. From a frequentist standpoint, overparameterization could be avoided by fixing the misclassification rate in a reasonable estimate for the sensitivity of the diagnostic test. A more elegant way of doing so is to use Bayesian analysis where the prior information about the misclassification rate takes the form of a probability distribution. However, both frequentist and Bayesian solutions show a main hurdle for their implementation in the research of ME/CFS. Given the lack of a disease biomarker, it is unclear which reasonable value or probability distribution to choose for the sensitivity of current diagnostic tools of ME/CFS.

As a final remark, our formulation of the misclassification problem assumed that misclassification is only dependent on the true clinical status of the suspected cases. In practice, it is very likely that misclassification is dependent on the symptoms’ severity profile of a given individual, or at least dependent on a given set of covariates. If so, Paulino et al. (Paulino et al., 2003) provided a Bayesian solution for modelling misclassification in this scenario. Given its technical complexity, we envision some difficulties in a wide application of this statistical solution by researchers of ME/CFS who are typically not trained in such advanced statistical methodology. To overcome this potential problem, we recommend a strong collaboration between these researchers and biostatisticians who have in principle the technical skills needed.

## Data Availability

The data under analysis is part of a database dedicated to Chronic Fatigue, UKMEB, and is the responsibility of the London School of Hygiene and Tropical Medicine working group.

## 7 Ethical approval

All participants provided written informed consent for data collection (questionnaire, clinical measurement and laboratory tests), and for allowing their samples to be available to any research receiving ethical approval. Participants received an extensive information sheet and consent form in which there was an option for participation withdraw from the study at any time. Ethical approval was granted by the LSHTM Ethics Committee (Ref. 6123) and the National Research Ethics Service (NRES) London-Bloomsbury Research Ethics Committee (REC ref. 11/10/1760, IRAS ID: 77765).

## Acknowledgements

The authors acknowledge the CureME group for providing the data from the UKMEB. JM acknowledges a PhD fellowship by the Fundação para a Ciência e Tecnologia, Portugal (ref. SFRH/BD/149758/2019).

## Notes

### Competing Interest Statement

The authors have declared no competing interest.

### Funding Statement

This work was partially funded by Fundação para a Ciência e a Tecnologia, Portugal (ref. SFRH/BD/149758/2019).

### Author Declarations

Ethical approval was granted by the LSHTM Ethics Committee (Ref. 6123) and the National Research Ethics Service (NRES) London-Bloomsbury Research Ethics Committee (REC ref. 11/10/1760, IRAS ID: 77765).

## References

Agresti, Alan. 2002. Categorical Data Analysis. John Wiley & Sons, Inc.

Brurberg, Kjetil Gundro, Marita Sporstøl Fønhus, Lillebeth Larun, Signe Flottorp, and Kirsti Malterud. 2014. “Case definitions for chronic fatigue syndrome/myalgic encephalomyelitis (CFS/ME): a systematic review.” BMJ Open 4:e003973.

Carruthers, Bruce M., Anil Kumar Jain, Kenny L. De Meirleir, Daniel L. Peterson, Nancy G. Klimas, A. Martin Lerner, Alison C. Bested, Pierre Flor-Henry, Pradip Joshi, A. C. Peter Powles, Jeffrey A. Sherkey, and Marjorie I. van de Sande. 2003. “Myalgic Encephalomyelitis/Chronic Fatigue Syndrome.” Journal of Chronic Fatigue Syndrome 11:7–115.

Carruthers, B. M., M. I. van de Sande, K. L. De Meirleir, N. G. Klimas, G. Broderick, T. Mitchell, D. Staines, A. C. P. Powles, N. Speight, R. Vallings, L. Bateman, B. Baumgarten-Austrheim, D. S. Bell, N. Carlo-Stella, J. Chia, A. Darragh, D. Jo, D. Lewis, A. R. Light, S. Marshall-Gradisbik, I. Mena, J. A. Mikovits, K. Miwa, M. Murovska, M. L. Pall, and S. Stevens. 2011. “Myalgic encephalomyelitis: International Consensus Criteria.” Journal of Internal Medicine 270:327–338.

Fukuda, Keiji, S. E. Strausm, I. Hickie, M. C. Sharpe, J. G. Dobbins, and A. Komaroff. 1994. “The Chronic Fatigue Syndrome: A Comprehensive Approach to Its Definition and Study.” Annals of Internal Medicine 121:953.

Gower, John C and Matthijs J Warrens. 2014. “Similarity, dissimilarity, and distance, measures of.” Wiley StatsRef: Statistics Reference Online pp. 1–11.

Institute of Medicine (US), Committee on the Diagnostic Criteria for Myalgic Encephalomyelitis/Chronic Fatigue Syndrome. 2015. Beyond Myalgic Encephalomyelitis/Chronic Fatigue Syndrome. National Academies Press.

Lacerda, Eliana M., Kathleen Mudie, Caroline C. Kingdon, Jack D. Butterworth, Shennae O’Boyle, and Luis Nacul. 2018. “The UK ME/CFS Biobank: A Disease-Specific Biobank for Advancing Clinical Research Into Myalgic Encephalomyelitis/Chronic Fatigue Syndrome.” Frontiers in Neurology 9.

Morris, Gerwyn and Michael Maes. 2013. “Myalgic encephalomyelitis/chronic fatigue syndrome and encephalomyelitis disseminata/multiple sclerosis show remarkable levels of similarity in phenomenology and neuroimmune characteristics.” BMC Medicine 11.

Nacul, Luís, Eliana M Lacerda, Caroline C Kingdon, Hayley Curran, and Erinna W Bowman. 2017. “How have selection bias and disease misclassification undermined the validity of myalgic encephalomyelitis/chronic fatigue syndrome studies?” Journal of Health Psychology 24:1765–1769.

Paulino, Carlos Daniel, Paulo Soares, and John Neuhaus. 2003. “Binomial Regression with Misclassification.” Biometrics 59:670–675.

Pheby, Derek F.H., Diana Araja, Uldis Berkis, Elenka Brenna, John Cullinan, Jean-Dominique de Korwin, Lara Gitto, Dyfrig A Hughes, Rachael M Hunter, Dominic Trepel, and Xia Wang-Steverding. 2020. “The Development of a Consistent Europe-Wide Approach to Investigating the Economic Impact of Myalgic Encephalomyelitis (ME/CFS): A Report from the European Network on ME/CFS (EUROMENE).” Healthcare 8:88.

Rasa, Santa, Zaiga Nora-Krukle, Nina Henning, Eva Eliassen, Evelina Shikova, Thomas Harrer, Carmen Scheibenbogen, Modra Murovska, and Bhupesh K. Prusty. 2018. “Chronic viral infections in myalgic encephalomyelitis/chronic fatigue syndrome (ME/CFS).” Journal of Translational Medicine 16.

Scheibenbogen, Carmen, Helma Freitag, Julià Blanco, Enrica Capelli, Eliana Lacerda, Jerome Authier, Mira Meeus, Jesus Castro Marrero, Zaiga Nora-Krukle, Elisa Oltra, et al. 2017. “The European ME/CFS Biomarker Landscape project: an initiative of the European network EUROMENE.” Journal of translational medicine 15:162.

